# Identifying Dimensions and Components of Organizational Readiness for Knowledge Translation Implementation in Type I Medical Sciences Universities in Iran: A Scoping Review

**DOI:** 10.1101/2025.06.30.25330171

**Authors:** Hamed Hosseinzadeh, Ali Sadatmoosavi, Oranus Tajedini, Asghar Tavan, Mohammad Azami, Mousa YaminFirooz

**Author notes:** Corresponding Author: Ali Sadatmoosavi ، Associate Professor, Department of Medical Library & Information Sciences, Faculty of Management and Medical Information Sciences, Kerman University of Medical Sciences., Kerman, Iran.

## Abstract

**Introduction:** Healthcare organizations must have the ability to adapt to rapid environmental changes and respond to evidence-based needs. One of the key prerequisites for the effective implementation of these changes is organizational readiness for knowledge translation (KT). The aim of this study is to identify the dimensions and components of organizational readiness for implementing KT in medical universities in Iran.

**Methods:** This research was conducted using a scoping review approach based on the Arksey and O’Malley framework. A comprehensive search was performed without a time limitation in international databases such as PubMed, Scopus, Web of Science, and Persian databases MagIran and SID. Out of 4540 identified documents, 14 articles were included after screening and final assessment.

**Results:** After extracting the dimensions and components from the reviewed studies, five dimensions and 14 components were identified, as follows: Organizational Climate (culture of readiness and innovation, intra- and inter-organizational interactions, organizational dynamics), Organizational Support (development and training opportunities, financial resources, capacity building and organizational performance improvement), Change Management (strategies and change implementation capacity, leadership support), Organizational Context (knowledge management in the organization, organizational structure and technological advancement, social capital), and Human Resources (self-efficacy and motivation, leadership, collaboration and participation).

**Conclusion:** To enhance organizational readiness for KT implementation, it is essential for health system managers and policymakers to understand the importance of utilizing research evidence and, through skill development and active participation in research processes, create the necessary environment for utilizing generated knowledge.

## Introduction

Healthcare organizations are complex and highly adaptable entities operating in a dynamic environment. Policymakers and healthcare managers must utilize available credible research evidence for planning, organizing, leading, and controlling healthcare organizations (1). Knowledge translation, policy-making, and management of healthcare organizations lead to optimal use of scarce resources and increased efficiency (2). Given the evidence that successful KT activities can significantly impact health outcomes, it is crucial to create evidence and strategies that support such activities in low- and middle-income countries (3). Such approaches can also facilitate knowledge exchange and influence evidence-based information uptake by transforming institutions in low- and middle-income countries into knowledge brokers (specialized experts who bridge the gap between research and end-users) (4). Factors related to capacity, interest, ability, and skills of both producers and users of knowledge and the organizational environment of healthcare systems affect the knowledge process (1).

Management experts argue that organizational readiness for change is a critical prerequisite for the successful implementation and coping with change. Many organizational innovations fail due to the lack of organizational readiness (5). Organizational readiness assessment is a method that evaluates various dimensions of an organization, such as managerial and organizational aspects, human resources, structural, process, technical, infrastructural, and cultural aspects, to assess the readiness of each part of the organization for adopting management systems. In fact, readiness is a necessary prerequisite for an individual or an organization to successfully confront organizational change (6). Research on organizational readiness for KT can inform capacity-building strategies that enhance the uptake and successful implementation of KT activities. This can improve the national credibility of the institution, increase future KT opportunities, and open doors for new partnerships and financial resources. This motivation for institutional change ultimately facilitates achieving the overarching goal of reducing disease burden and mortality, which is one of the key objectives of KT activities (3).

One of the concerns in the global healthcare system is the lack of credible evidence and research-derived knowledge (7). Despite the exponential growth in publications related to the importance of KT across different health sciences, there is still a significant gap between what is being done (practice) and what is known (evidence) at all levels of healthcare decision-making (8). In Iran, despite significant growth in domestic research in the health system field in recent years, research is still in its infancy. As a result, policymakers often face a lack of reliable evidence when addressing many questions (9). Since one of the challenges of the health system is the need for research to support policy-making, it is necessary to develop applied research in the health system, especially in the field of KT (10). Considering these issues and their importance, this research aims to identify the dimensions and components of organizational readiness for implementing KT in medical universities in Iran.

## Materials and Methods

This study was conducted using a scoping review approach based on the five-stage framework proposed by Arksey and O’Malley (2005). The stages include: 1) Formulation of the research question, 2) Identification of relevant studies, 3) Selection of eligible studies, 4) Extraction and categorization of data, and 5) Analysis, synthesis, and presentation of results [11]. This approach was chosen to comprehensively and systematically explore key concepts in the studied area, particularly in emerging and complex topics such as organizational readiness for KT implementation.

Scoping reviews enable the integration and analysis of a wide range of published and unpublished studies, providing a broad view of the various dimensions of a phenomenon, especially when the goal is to go beyond measuring the effectiveness of an intervention and focus on elucidating frameworks and conceptual models [12].

### Stage 1: Formulation of the Research Question

The main research question was: “What are the dimensions and components of organizational readiness in Iranian medical universities for the effective implementation of knowledge translation?”

### Stage 2: Identification of Relevant Studies

No time limitations were set for the search. Electronic searches were conducted in credible international databases, including PubMed, Scopus, Web of Science, and Google Scholar. For Persian articles, MagIran and SID databases were reviewed. The key search terms included: Translation of knowledge, use of research, knowledge transfer, knowledge exchange, knowledge into practice, knowledge mobilization, organizational readiness, organization and reading, organization and readiness In Persian-language databases, equivalent MeSH terms in Farsi were used accordingly (see Table 1).

**Table 1:**
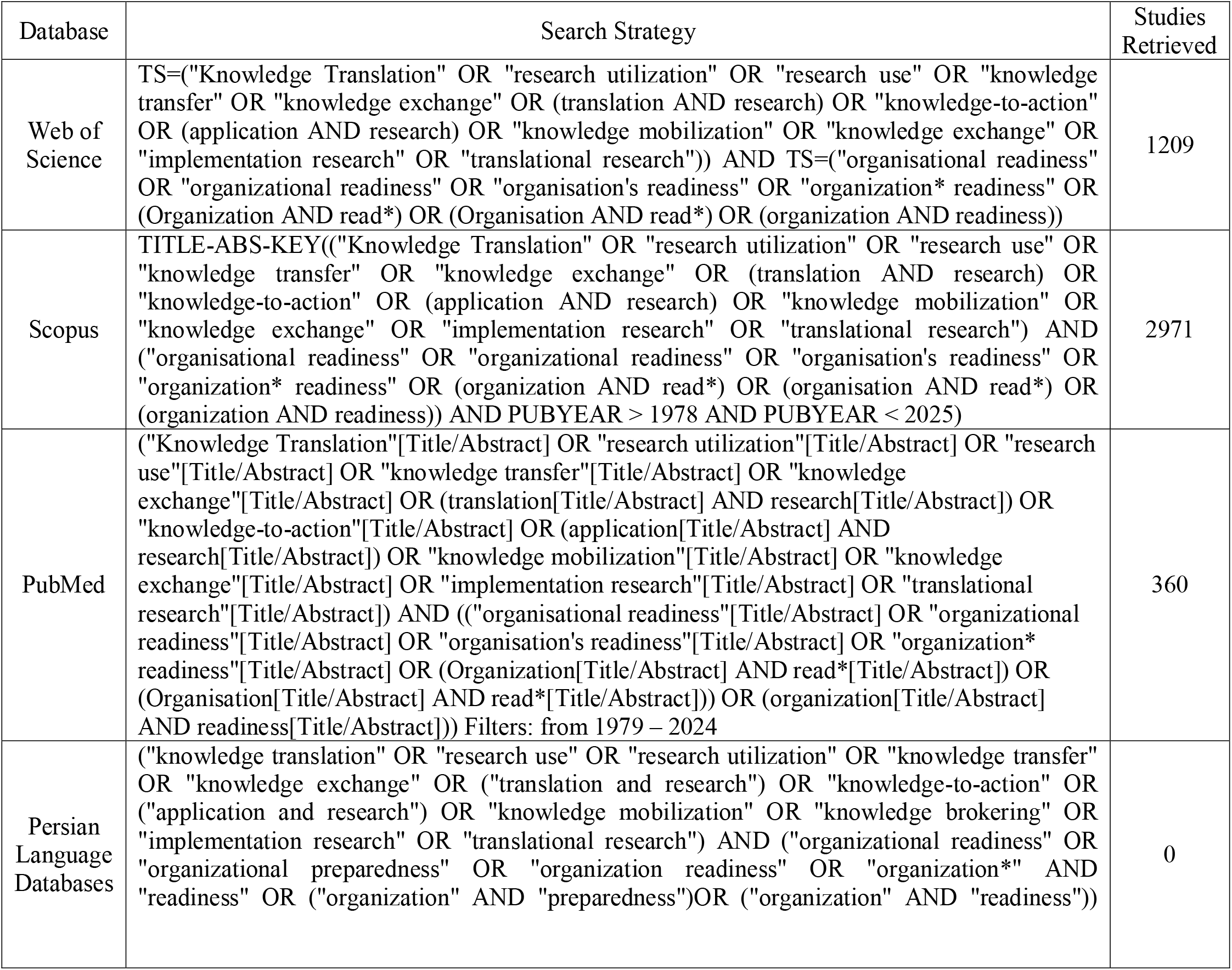
Search Strategy of Databases and Number of Studies Retrieved from the Reviewed Databases.

Inclusion criteria were: 1) Articles in Persian and English, and 2) Studies related to organizational readiness and KT in health. Exclusion criteria were: 1) Irrelevant articles, and 2) Articles published in languages other than Persian and English.

### Stage 3: Selection of Eligible Studies

A total of 4540 articles were retrieved from the mentioned databases. Data were managed using EndNote software, and after removing duplicates (n=1674), 2866 articles were left for title and abstract screening. Subsequently, 2819 articles were excluded due to content mismatch, and 47 articles were selected for full-text review. Finally, 14 articles were included in the analysis (The PRISMA-ScR flowchart is presented in Figure 1).

**Figure 1:**
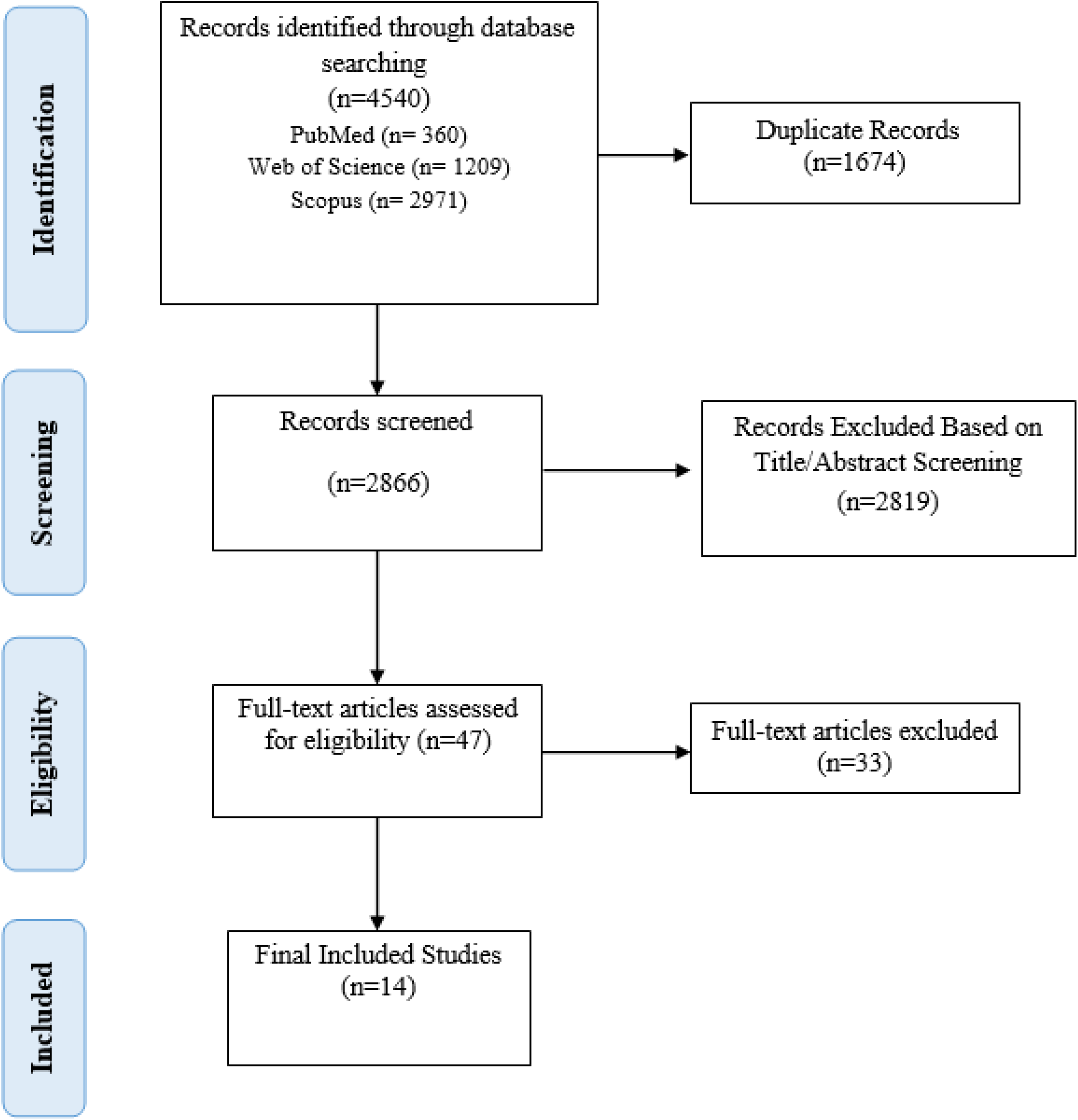
PRISMA Flow Diagram.

**Figure 2:**
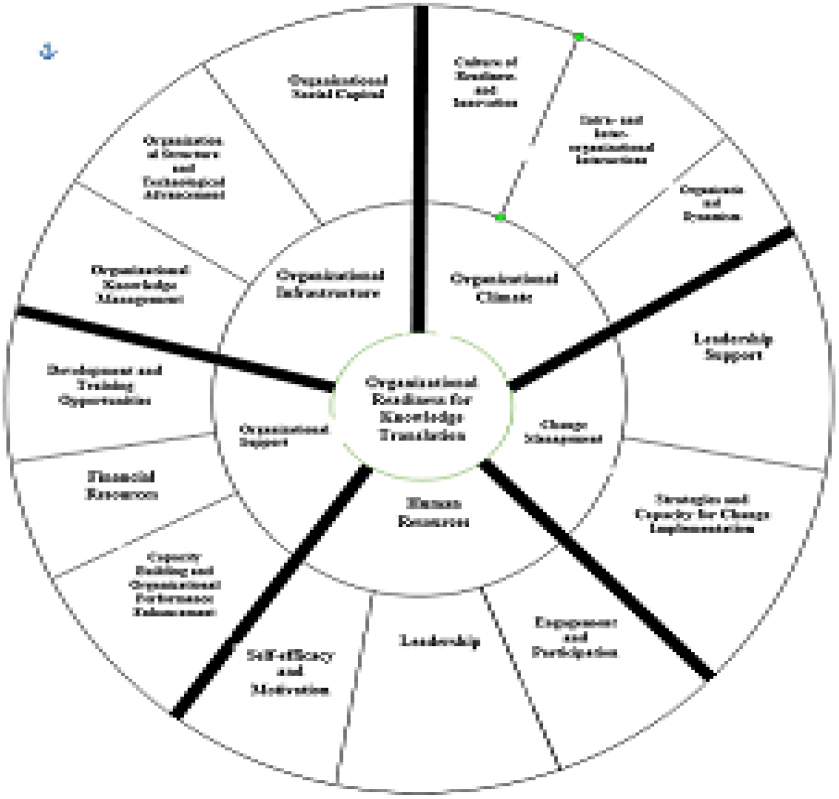
Conceptual Model of Organizational Readiness for Implementing Knowledge Translation.

### Stage 4: Extraction and Categorization of Data

The key data from the selected studies were summarized and categorized in Table 2. These data included study objectives, methodologies, tools used, and the dimensions and components identified regarding organizational readiness for KT.

**Table 2.** Final list of studies with detailed description of each study.

| Researchers | Year | Location | Study Type | Tool | Topic | Summary of Findings |
| --- | --- | --- | --- | --- | --- | --- |
| Kalbarczyk, A., et al. | 2024 | Bangladesh, Ethiopia, DRC, India, Indonesia, Nigeria | Quantitative | Survey checklist | Factors affecting institutional readiness for KT in LMIC academic institutions | Readiness assessment can be used to systematically identify needs and develop targeted strategies. |
| Guleid, F. H., et al. | 2022 | Kenya | Qualitative | Interview | Experience of Kenyan researchers and policymakers with KT during COVID-19 | This study provides a real-world example of KT use during a public health crisis. |
| Kalbarczyk, A., et al. | 2021 | Bangladesh, Ethiopia, DRC, India, Indonesia, Nigeria | Mixed-methods | Questionnaire | Developing a tool to assess institutional readiness for KT in LMICs | The study identifies a tool and unique opportunities for capacity building in LMIC universities. |
| Kalbarczyk, A., et al. | 2021 | Bangladesh, Ethiopia, DRC, India, Indonesia, Nigeria | Qualitative | Interview | Barriers and facilitators to KT in academic institutions in LMICs | Cross-cutting themes highlighting barriers and enablers can inform readiness-building strategies. |
| Gagnon, M. P., et al. | 2019 | Spain and Canada | Commentary | Correspondence | Future directions for the OR4KT readiness tool | Describes the development, cross-cultural adaptation, and content validation of the OR4KT tool. |
| Lisa M. Puchalski Ritchie | 2019 |  | Commentary |  | Organizational readiness for change in KT tool evaluation | Highlights the strengths of OR4KT including its foundation on systematic reviews. |
| Gagnon, M. P., et al. | 2018 | Spain and Canada | Review | Questionnaire | Development and validation of a cross-cultural OR4KT readiness assessment tool | OR4KT is available as a KT readiness measure in healthcare organizations. |
| Roberto Nuño-Solinís | 2018 |  | Commentary |  | Organizational readiness for change in healthcare | OR4KT helps organizations assess readiness across six dimensions. |
| Grandes, G., et al. | 2017 | Spain | Quantitative | Questionnaire | Validation of the Spanish version of OR4KT | Spanish version of OR4KT demonstrated strong reliability and validity. |
| Gagliardi, A. R. and M. J. Dobrow | 2016 | Canada | Qualitative | Interview | Conditions needed for integrated knowledge translation (IKT) | Use of IKT framework may help more organizations become ready for IKT. |
| Attieh, R., et al. | 2014 | Canada | Review | Questionnaire | KT readiness in chronic care: A Delphi study | Study identified key dimensions and subdimensions for assessing KT readiness. |
| Gagnon, M. P., et al. | 2014 | Canada | Mixed-methods | Checklist | Systematic review of tools to assess organizational readiness for KT in healthcare | Few validated and reliable tools for KT readiness in healthcare exist. |
| Attieh, R., et al. | 2013 | Canada | Review | Checklist | Theoretical components of KT readiness in chronic care | Five main concepts identified: organizational dynamics, change process, innovation readiness, institutional readiness, personal readiness. |
| Gagnon, M. P., et al. | 2011 | Canada | Review | Questionnaire & checklist | Measuring organizational readiness for KT in chronic care | Comprehensive synthesis of explanatory KT models and tools. |

**Table 3:** Dimensions and Components Extracted from the Reviewed Articles.

| Researchers and Year | Subject | Study Type | Dimension | Components |
| --- | --- | --- | --- | --- |
| Kalbarczyk et al., 2021; Gagnon et al., 2018; Attieh et al., 2014 | Factors influencing KT; tools for evaluating readiness | Quantitative, Mixed-method, Qualitative | Organizational Climate | Organizational culture and values, support for KT, recognition and rewards |
| Grandes et al., 2017; Nuño-Solinís, 2018 | Assessing readiness for change | Quantitative, Interpretive | Organizational Climate | Organizational openness to change, staff cohesion, internal communication, job stress |
| Guleid et al., 2022; Gagnon et al., 2019 | Knowledge translation in COVID-19; OR4KT tool | Qualitative, Mixed-method, Interpretive | Effectiveness of Organizational Change | Organizational capacity to initiate and sustain change, leadership support, change in management strategy |
| Kalbarczyk et al., 2021; Attieh et al., 2014 | External motivations and pressures to implement KT | Qualitative, Mixed-method | Prioritization and Global Alignment | Alignment of KT with organizational priorities; engagement with international actors |
| Kalbarczyk et al., 2021 | Individual barriers to KT implementation | Qualitative, Quantitative | Self-efficacy | Individuals' belief in their ability to effectively participate in KT; communication skills; adaptability to challenges |
| Kalbarczyk et al., 2021; Nuño-Solinís, 2018 | Infrastructure supporting KT | Mixed-method, Interpretive | Financial and Human Resources | Access to funding, human resources, financial sustainability; allocation of resources for training |
| Guleid et al., 2022 | Policymaker experiences during the pandemic | Qualitative | Institutionalization of KT | Developing KT strategies and policies, establishing structures, evaluation mechanisms, capacity building |
| Kalbarczyk et al., 2021;<br>Gagliardi & Dobrow,<br>2016 | Facilitating<br>evidence exchange | Qualitative | Networking and<br>Communication | Collaboration between researchers<br>and policymakers, active<br>participation in policymaking |
| Gagnon et al., 2018;<br>Ritchie, 2019; Attieh et<br>al., 2014 | Role of leadership<br>in enhancing KT | Interpretive<br>, Mixed-method,<br>Review | Leadership and<br>Organizational<br>Support | Leadership style, leader<br>participation, organizational<br>support, governance structures |
| Gagnon et al., 2019;<br>Nuño-Solinís, 2018 | Motivation to<br>participate in KT<br>activities | Qualitative,<br>Interpretive | Staff Motivation<br>and Commitment | Individual and collective<br>motivation, incentives, goal clarity |
| Gagnon et al., 2018;<br>Kalbarczyk et al., 2021 | Readiness in<br>different contexts | Review,<br>Interpretive | Organizational<br>Context | Environmental factors,<br>infrastructure, supportive policies,<br>organizational size and complexity |
| Nuño-Solinís, 2018;<br>Ritchie, 2019 | Evaluation of<br>change programs | Review,<br>Interpretive | Content and<br>Complexity of<br>Change | Characteristics of proposed<br>change, supporting evidence,<br>complexity and adaptability |
| Guleid et al., 2022;<br>Gagliardi & Dobrow,<br>2016 | Developing KT<br>capabilities | Qualitative | Capacity<br>Building and<br>Training | Training researchers and<br>policymakers, skill enhancement |
| Guleid et al., 2022;<br>Kalbarczyk et al., 2021 | Responding to<br>crises | Qualitative | Timing and<br>External<br>Pressures | Timely evidence production,<br>urgent policy needs, peer pressure |

### Stage 5: Analysis and Presentation of Results

Based on content analysis of the selected studies, five main dimensions and fourteen components were identified and extracted as a conceptual framework for organizational readiness to implement KT in Iranian medical universities (Figure 1).

## Results

Analysis of the results from the search indicated that the majority of the retrieved articles came from the Scopus database (2971 articles), followed by PubMed (360 articles) and Web of Science (1209 articles). However, in Persian-language databases like MagIran and SID, due to the novelty of the topic and limited relevant studies, no articles met the inclusion criteria (Flowchart).

It is noteworthy that a significant portion of the retrieved articles were focused on two separate domains: “Organizational Readiness” and “Knowledge Translation,” with only a few addressing the relationship between the two. After final screening, 14 relevant articles were selected for final analysis.

## Discussion and Conclusion

The findings of this scoping review study revealed that organizational readiness for implementing knowledge translation in medical universities in Iran consists of five key dimensions: organizational climate, organizational support, change management, organizational context, and human resources, which can be analyzed through 14 subcomponents. These dimensions and components are directly related to the organization’s ability to plan, make decisions, and effectively execute knowledge translation activities.

**Organizational climate** has been identified as one of the key dimensions of organizational readiness for implementing knowledge translation in 7 out of the 14 reviewed studies. This dimension refers to the organizational environment and culture, which can either support or hinder knowledge translation activities [13]. According to the findings of the reviewed studies, the components of this dimension include readiness culture, readiness for innovation, intra- and inter-organizational interactions, and organizational dynamics.

The study by Kalbarczyk et al. (2024) showed that, in the context of knowledge translation readiness, organizational climate (at the organizational level) is more relevant for younger individuals, while self-efficacy (at the individual level) is more significant for middle-aged and older groups [13]. This finding suggests that less experienced individuals may rely more on internal support for knowledge translation, as they do not yet have sufficient external networks and resources. In contrast, more experienced individuals, with greater mastery, feel more control over their environment and have greater confidence in their ability to implement knowledge translation.

Other studies have also emphasized the importance of organizational climate in supporting organizational dynamics. For example, the findings of Grandes et al. (2017) and Attieh et al. (2014) suggest that a favorable organizational climate facilitates positive organizational changes, strengthens trust among employees, and promotes collaboration between them [14,15].

The results of several other studies also confirm the impact of organizational climate on the knowledge-sharing process within organizations. Among these studies are those by Akram et al. (2020) [16], Fatemi et al. (2022) [17], Kaffashan Kakhki et al. (2020) [18], Mbandi (2020) [19], Usmanova et al. (2020) [20], and Vandavasi et al. (2020) [21]. The findings of these studies indicate that organizational climate can either facilitate or hinder knowledge interactions among employees.

For instance, the study by Kuo et al. (2014) showed that a supportive organizational climate increases employees’ intrinsic motivation to share and receive knowledge, while a negative perception of organizational culture prevents knowledge exchange among colleagues [22]. Additionally, Betes and Khasawneh (2005) confirmed that organizational climate influences employees’ perceptions of the prevailing culture [23]. The findings of Banerjee et al. (2017) also showed that a positive organizational climate can facilitate knowledge sharing among employees [24]. Overall, organizational climate can be seen as a soft yet effective foundation for the success of knowledge translation. Medical organizations should strengthen their organizational readiness by creating an innovation culture, fostering constructive interactions between units, and enhancing organizational dynamics to ensure the effective implementation of knowledge translation.

**In the present review, organizational support** was identified as one of the key dimensions of readiness for implementing knowledge translation in 7 out of the 14 reviewed studies. This dimension includes three main components: development and training opportunities, financial resources, and capacity building and performance improvement. Organizational support plays a fundamental role in creating the necessary infrastructure and providing the conditions required for the establishment and sustainability of knowledge translation in medical organizations. The study by Attieh et al. (2014) showed that this dimension is achieved through processes such as feedback, evaluation, monitoring, and creating a supportive climate. Moreover, the successful implementation of any organizational change requires an internal environment that can stabilize and institutionalize that change [15]. Our findings also emphasize that organizational support encompasses a range of requirements, including access to financial resources, supportive policies and procedures, training opportunities, and institutional and individual readiness. According to the study by Kalbarczyk et al. (2024), financial resources played a lesser role in the perception of readiness for knowledge translation among individuals in coordination roles. This finding may be due to the lesser involvement of these individuals in budgeting processes and financial resource management [13]. On the other hand, the study by Attieh et al. (2014) introduced the concept of “institutional readiness,” which consists of motivation, values, organizational goals, and managerial and structural support [15]. In this context, systematic readiness assessment can help identify gaps and design targeted strategies for enhancing knowledge translation. Studies conducted in Iran have also confirmed the importance of this dimension. Specifically, the research by Shams and Abbasi-Kasani (2017), Ghanbari et al. (2017), Lotfi et al. (2016), Diba Vajari et al. (2016), and Youzbashi et al. (2016) showed that managerial support was the most important facilitating factor in the successful transfer of education and the implementation of knowledge programs in healthcare organizations. These findings align with the results of studies by Kontoghiorghes (2014), Lancaster (2016), and Park et al. (2018), which highlighted the role of structural and managerial support in the success of knowledge translation [25-32]. Overall, the dimension of organizational support, by providing resources, structures, learning opportunities, and capacity building, forms the essential infrastructure required for the readiness and successful implementation of knowledge translation.

**In this scoping review, change management** was identified in 7 out of the 14 reviewed studies as one of the key dimensions of organizational readiness for implementing knowledge translation (KT). This dimension refers to the organization’s ability to plan, lead, execute, and institutionalize changes required for the successful implementation of KT activities. Accordingly, organizational members share beliefs about their collective ability to manage and implement change [13]. Findings from this review indicate that change management includes two main components: capacity to implement organizational change, leadership support for change, and shifts in managerial strategies.

Kalbarczyk et al. (2021) emphasized that assessing organizational readiness can serve as an effective tool for identifying facilitators and barriers to implementing capacity-building strategies. This assessment can also enable organizations to tailor their approaches to overcome obstacles in implementing KT [3]. The study by Kalbarczyk et al. (2024) further highlighted the role of leadership and gender in the effectiveness of organizational change. Their findings indicate that active engagement of organizational leaders and promotion of women’s leadership can enhance the effectiveness of KT-related changes [13].

Strauss et al. (2009) reported that the effectiveness of change at the organizational level contributed more significantly to readiness for KT among men compared to women. Additionally, for individuals in leadership roles, the effectiveness of change was less significant than for those in other professional roles. Nevertheless, supportive leadership, while not sufficient on its own, is recognized as an important and well-documented component of the organizational change management process [25].

In line with this, the study by Shea et al. (2014) evaluated the psychometric properties of organizational readiness assessment tools, including the change effectiveness scale. In these tools, change effectiveness was primarily based on trust in the ability to manage the change process, coordination of tasks, and organizational investment motivation. The related items in the present study similarly describe elements of trust, motivation, and readiness for change [26].

From a theoretical standpoint, organizational members may commit to change for different reasons—some may perceive it as valuable, while others may feel obliged or responsible. It is reasonable to assume that the perspectives and actions of external stakeholders may also influence these values and commitments, particularly in health-related institutions [3]. The concept of the “change process” refers to the set of actions pursued to effectively implement organizational change. This process includes managerial support, external influences, and perceived options for change.

According to the model proposed by Armenakis and Harris (2002), this process consists of three stages: readiness, adoption, and institutionalization. Since these stages do not occur automatically, managers must design and implement intelligent, capacity-aligned programs to foster ongoing readiness for change among staff members [27]. Complementing this framework, Armenakis et al. (2011) defined readiness for change as a combination of beliefs, attitudes, and behavioral intentions that reflect both individual and organizational perceptions of the feasibility of successful change [28].

Therefore, the success of medical universities in implementing knowledge translation requires simultaneous attention to change strategies, the capacity to execute them, and supportive leadership

**In the present scoping review, the dimension of *organizational context*** was identified in 7 out of the 14 final studies as one of the core components of organizational readiness for knowledge translation (KT). This dimension refers to the capacities, infrastructures, and contextual features of an organization that either facilitate or hinder the effective implementation of KT. Various levels of organizational context involve skill and training deficiencies among researchers and policymakers, weaknesses in institutional infrastructure, and insufficient human and technical resources [3].

Gagnon et al. (2019) demonstrated that assessing the organizational context—including resources, infrastructure, and external factors—can play a crucial role in the success or failure of KT processes [29]. Similarly, Kalbarczyk et al. (2021) emphasized that organizational structures such as physical spaces, specialized training, and adequate human resources are essential prerequisites for supporting KT activities [3]. According to our findings, the organizational context dimension includes four key components: knowledge management, organizational structure, technological advancement, and organizational social capital.

The study by Gagnon et al. (2011) showed the direct impact of organizational context on research utilization and highlighted the need for health systems to be adequately prepared to apply scientific evidence. Facilitating factors in this process include strong leadership, participatory decision-making, financial and institutional support, effective infrastructure, and dynamic organizational communication [30].

Kalbarczyk et al. (2021) also identified soft skills (e.g., communication, self-awareness, adaptability), robust professional networks, and alignment of organizational priorities and incentives as important drivers of KT capacity building in low- and middle-income countries (LMICs) [3]. In this regard, the capacity-building model proposed by Potter and Brough (2004) introduces interrelated and hierarchical dimensions including structures, systems, roles, staff, infrastructure, skills, and tools. According to this model, robust and systematic structures enable the optimal use of tools and skills to facilitate effective change [31].

El-Jardali and Fadlallah, in their KT framework, placed researcher capacity-building at the core and stressed that interventions should simultaneously target individual, team, institutional, and system levels [32]. Although institutional capacity is widely acknowledged as a prerequisite for the sustainable use of evidence, there is limited evidence on how to effectively implement capacity-building activities at this level [33].

In this framework, Attieh et al. (2014) categorized contextual organizational factors into two main subsets: appropriate human resources and sufficient material resources to support change, which were considered more critical than organizational culture [15]. Numerous other studies—including those by Weiner, Holt, and Kitson—have emphasized that without understanding and enhancing the organizational context, successful implementation of organizational changes, including KT, is unlikely [5, 34, 35]. These factors include adequate human and material resources, as well as an understanding of the prevailing organizational culture and climate in which change takes place [34, 36].

It is worth noting that capacity-building must align with the stage of organizational change, and strategies should be designed to facilitate the organization’s growth and evolution according to the readiness level and needs at each phase of change [13]. Finally, Murunga et al. (2020) highlighted the need for further research into the facilitators and barriers affecting researchers’ ability to successfully implement KT activities at both individual and organizational levels in LMICs [37].

In conclusion, it appears that medical science organizations must focus on the three main components of organizational context—knowledge management, structure and technology, and social capital—and simultaneously consider both internal and external factors to create the necessary conditions for effective KT implementation.

**The *human resources* dimension** was identified in 12 of the final included studies and is regarded as one of the most fundamental strategic assets of an organization. According to the findings of this review, the key components of this dimension include self-efficacy, leadership, motivation, interaction, and participation. *Self-efficacy* refers to individuals’ belief in their capability to contribute effectively to KT activities and plays a central role in personal readiness for engaging in organizational change processes (13, 41). The study by Kalbarczyk et al. (2021) also indicated that lack of training and necessary resources—such as time and personnel—constitutes a major challenge for KT implementation, and stressed the necessity of leadership engagement to strengthen KT efforts (3).

Organizational leaders, through consistent messaging and action, can foster a shared vision for change among staff and help enhance motivation and engagement (38). Leadership traits that support change—such as high self-efficacy—are recognized as facilitators in creating an enabling environment for early-career researchers (39, 40). Furthermore, self-efficacy develops in interaction with the organizational context and grows through repeated mastery experiences and modeling (42). It is expected that more experienced staff, possessing higher levels of self-efficacy, will have greater confidence in their ability to implement KT.

Numerous studies have also confirmed the role of participatory leadership and fair treatment of employees in enhancing organizational trust, motivation, and effective engagement (43, 44). For instance, Buchan et al. (2005) found that involving staff in decision-making—even in the face of increased workload—leads to improved morale and a greater sense of unity within the organization (43). Similarly, Liu’s (2008) research highlighted the importance of interpersonal relationships and effective communication between managers and staff in the success of participatory management (45).

Despite the emphasis placed on human resources by various studies, some findings did not align with this perspective. Notably, Chang & Huang (2005) found that strategic human resource management does not necessarily have a significant impact on organizational performance, and no single HR management approach proved superior to others (46).

Overall, this scoping review reveals that human resources play a critical role in strengthening organizational capacity for implementing KT. To achieve effective readiness, medical universities should simultaneously focus on enhancing individual self-efficacy, developing participatory leadership, fostering staff motivation, improving interpersonal interactions, and engaging stakeholders.

These findings demonstrate that *organizational readiness for knowledge translation* is a complex and multidimensional phenomenon influenced by a combination of organizational and individual factors, including organizational climate, institutional support, change management, organizational context, and human resources. In alignment with international studies, these dimensions underscore that healthcare and academic organizations must pay strategic attention to such factors to successfully implement evidence-based change initiatives.

Based on the results of this study, a conceptual framework has been proposed for assessing organizational readiness to implement KT within Iranian medical universities. This framework may serve as a strategic tool for managers, planners, and health policymakers to focus on the identified key components and thus create the necessary conditions for effective KT processes. Although the core dimensions of organizational readiness are generally consistent across studies, the prioritization and weighting of each dimension vary depending on the cultural, social, and economic context. Therefore, integrating globally recognized indicators with locally adapted components can provide a comprehensive evaluation framework for higher education institutions in the health sector in Iran.

Furthermore, strengthening transformational leadership, promoting a culture of innovation, expanding inter-organizational collaboration, and developing organizational change capacity require targeted investment and fundamental revisions in traditional academic and research structures. In this regard, *medical librarians* can play a pivotal role as knowledge intermediaries, evidence exchange facilitators, and advisors in clinical and educational decision-making. Ultimately, the conceptual model derived from this study can inform the development of practical and localized assessment tools in the KT domain and serve as an operational guide for policymakers and decision-makers in Iran’s health and medical education systems

**The limitations** of this study included restricted access to certain international databases due to sanctions (such as Scopus), which may have affected the comprehensiveness of the literature review. Additionally, the focus on type one medical universities limits the generalizability of the findings to other institutions and requires cautious interpretation. Among the strengths of this study were the systematic identification of key dimensions and components of organizational readiness for knowledge translation, which provided both a theoretical and operational framework for policymakers and educational managers. Furthermore, the integration of qualitative and quantitative methods contributed to a deeper extraction of concepts and enhanced the credibility of the findings.

## Data Availability

All relevant data are within the paper however, any question or other file data is required you can contact us using the email address, upon reasonable request.

## Acknowledgments

we thanks to clinical research development unit of rohani hospital.

## Funding

The author(s) received no financial support for the research, authorship, and/or publication of this article

## Declarations

## Ethics approval and consent to participate

This survey was carried out in accordance with the guidelines of the Declaration of Helsinki, was approved by the Ethics Committee of Kerman University of Medical Sciences (Ethics Code: IR.KMU.REC.1402.312).

## Consent for publication

Not applicable.

## Competing interests

The authors declare no competing interests

## Notes

### Competing Interest Statement

The authors have declared no competing interest.

### Funding Statement

This study did not receive any funding

### Summary of Updates

Introduction: Healthcare organizations must have the ability to adapt to rapid environmental changes and respond to evidence-based needs. One of the key prerequisites for the effective implementation of these changes is organizational readiness for knowledge translation (KT). The aim of this study is to identify the dimensions and components of organizational readiness for implementing KT in medical universities in Iran. Methods: This research was conducted using a scoping review approach based on the Arksey and O'Malley framework. A comprehensive search was performed without a time limitation in international databases such as PubMed, Scopus, Web of Science, and Persian databases MagIran and SID. Out of 4540 identified documents, 14 articles were included after screening and final assessment. Results: After extracting the dimensions and components from the reviewed studies, five dimensions and 14 components were identified, as follows: Organizational Climate (culture of readiness and innovation, intra- and inter-organizational interactions, organizational dynamics), Organizational Support (development and training opportunities, financial resources, capacity building and organizational performance improvement), Change Management (strategies and change implementation capacity, leadership support), Organizational Context (knowledge management in the organization, organizational structure and technological advancement, social capital), and Human Resources (self-efficacy and motivation, leadership, collaboration and participation). Conclusion: To enhance organizational readiness for KT implementation, it is essential for health system managers and policymakers to understand the importance of utilizing research evidence and, through skill development and active participation in research processes, create the necessary environment for utilizing generated knowledge.

